# Direct comparison of venipuncture serum draws versus whole blood finger-stick specimens by anti-COVID-19 IgG/IgM rapid lateral flow immunoassay and ELISA

**DOI:** 10.1101/2021.06.04.21258182

**Authors:** Irfan Baig, Christian Tagwerker, Eric J. Brunson, Kristine Mundo, Davan Dutra-Smith, Mary Jane Carias-Marines, Ranulu Samanthi de Zoysa, David J. Smith

## Abstract

During the COVID-19 pandemic, manufacturers have developed several diagnostic test kits that include lateral flow immunoassays (LFIA) also known as rapid cassette testing. Rapid cassette testing provides qualitative test results indicating the presence or absence of IgG and IgM antibodies to determine COVID-19 (SARS-CoV-2) infection among individuals. Venipuncture blood draws have been the traditional and widely proposed sample collection method but is costly and not applicable to point-of-care testing (POC) and in remote settings. Whole blood finger-stick blood collections traditionally used by diabetics for glucose level testing is an ideal scenario, but raises concerns regarding the outcome of test results in regards to specificity and sensitivity. In this study we directly compare simultaneous collections of venipuncture serum (SST) blood draws and whole blood finger-sticks (n = 75) to detect human Anti-COVID-19 IgG and IgM antibodies using an EUA-approved lateral flow immunoassay, showing equal to enhanced performance characteristics for this specimen type.

## 1 Introduction

The SARS-CoV-2 (severe acute respiratory syndrome coronavirus 2 or COVID-19) has affected the global population with high transmission and infection rates [1]. SARS-CoV-2 is a single-stranded RNA coronavirus [2]. Comparisons of the genetic sequences of this virus have shown similarities to SARS-CoV and bat coronaviruses [3]. In humans, coronaviruses cause respiratory infections and are composed of several proteins including the spike (S), envelope (E), membrane (M), and nucleocapsid (N) protein [4], [5]. An in-depth study concluded that the S-protein holds enough affinity to the Angiotensin converting enzyme 2 (ACE2) receptor and utilizes it as a cell entry mechanism after human transmission of coronaviruses, which is largely thought to occur among close interactions via respiratory droplets produced by sneezing and coughing [6]. High titers of immunoglobulin IgG and IgM can be created in reaction to COVID-19 antigens and are sustained in the body after early contact for extended periods [7]. COVID-19 control was successful in countries where a combination of lockdown, quarantine, social distancing measures, and mass testing was performed [8], [9]. Several COVID-19 serologic rapid diagnostic tests were made available in early 2020 in form of lateral flow immunoassay (LFIA) cassettes [10], [11]. The COVID-19 IgG/IgM Rapid Test Cassette from Healgen® was tested on 04/21/2020 at the Frederick National Laboratory for Cancer Research (FNLCR), a Federally Funded Research and Development Center (FFRDC) sponsored by the National Cancer Institute (NCI). The COVID-19 IgG/IgM Rapid Test Cassette is intended to qualitatively detect IgM and IgG separately. As of May 29, 2020, the U.S. Food and Drug Administration (FDA) provided Emergency Use Authorization (EUA) for the Healgen® COVID-19 IgG/IgM rapid test cassette serology test in human venous whole blood, plasma from anticoagulated blood (Li+ heparin, K2EDTA and sodium citrate), or serum [12], [13]. The following study compares whole blood finger-stick collections applied to the rapid cassette from Healgen® versus simultaneous venipuncture serum collections verified by the EDI™ Novel Coronavirus COVID-19 IgG/IgM ELISA (enzyme-linked immunosorbent assay) test kit to show COVID-19 IgG/IgM antibodies can be detected reliably by whole blood finger-stick collection in combination with this rapid, lateral flow immunoassay cassette in remote (point-of-care) settings.

## 2 Materials and Methods

### 2.1 Study population

This study was approved by the Alcala Pharmaceutical Inc. Institutional Review Board (IORG0010127) in consideration of the Code of Ethics of the World Medical Association (Declaration of Helsinki). All collection activities occurred at the phlebotomy collection site at Alcala Testing and Analysis Services or the San Diego Comprehensive Pain Management Clinic (SDCPMC), 3703 Camino del Rio South, San Diego, CA, 92108 by certified phlebotomists or licensed vocational nurses. Serum ELISA testing and comparison to rapid cassette results were conducted from 06/01/2020 to 07/27/2020 at Alcala Testing and Analysis Services. All subjects were recruited via phone or e-mail announcements with phone follow-up or by word of mouth at the San Diego Comprehensive Pain Management Clinic, 3703 Camino del Rio South, San Diego, CA, 92108. Healthcare workers from the clinic also volunteered. Healthcare workers or volunteers in any setting could enroll without a prior COVID-19 test. All specimens derived from human subjects were de-identified of their health information as defined by Health Insurance Portability and Accountability Act (HIPAA) guidelines. Seventy-five volunteers took part in a whole blood finger-stick collection with immediate application to the Healgen® COVID-19 IgG and IgM antibody rapid cassette and a simultaneous venipuncture draw of serum (SST tiger top tube) analyzed by the EDI™ Novel Coronavirus COVID-19 IgG/IgM ELISA test procedure.

### 2.2 Test Instruments

Safire2 spectrophotometric microplate reader (TECAN, Austria), microplate strip washer (BIOTEK ELx-50, Winooski, VT), incubating microplate shaker, temperature controlled at ambient +5 to 65°C (TALBOYS, Thorofare, NJ). Bioshake XP 96-well plate shaker and ZR-96 MagStand or Ambion AM10027 magnetic plate stand. Eppendorf PCR chiller plate. Biorad® C1000 Touch™ Thermal Cycler. Luminex® MAGPIX® RT-PCR instrument.

### 2.3 Materials, collection and test kit components

EDI™ Novel Coronavirus COVID-19 IgG/IgM ELISA kits were purchased from Epitope Diagnostics Inc. (San Diego, CA) and stored at 2–8°C upon receipt. Healgen® lateral flow rapid cassettes were purchased from Healgen® Scientific LLC (Houston, TX) and stored at 2 – 30°C. For serum (SST) venipuncture draws standard phlebotomy vacutainer SST tubes and materials were purchased from Becton, Dickinson and Company (BD, Franklin Lakes, NJ). For whole blood finger-stick collections a kit including 21G (2.2. mm depth) high flow safety lancets (One-Care®, Irvine, CA), alcohol prep pad, gauze and band-aid were used. Digital timers ensured rapid cassette image interpretation at the recommended 10-minute mark. NxTAG® CoV Extended Panel Assay kits were purchased from Luminex® (Austin, TX) and stored at 2–8°C. Quick-DNA/RNA Viral MagBead™ Viral Kits R2140/R2141 were purchased from ZymoResearch (Irvine, CA) and stored at room temperature.

### 2.4 Sample processing and storage

At least 20 μL of human serum (collected in serum SST tubes by venipuncture) was used for measurement in duplicate using EDI™ Novel Coronavirus COVID-19 IgG/IgM ELISA test kits. Samples were processed on the same day. Severe hemolytic samples were excluded. Healgen® lateral flow rapid cassette kits for COVID-19 IgG and IgM antibody detection were used for testing individuals by whole blood finger-stick at the Alcala Labs collection site or San Diego Comprehensive Pain Management Clinic by a certified phlebotomist/nurse. Simultaneous serum SST venipuncture draws were carried out with every whole blood finger-stick. For 8 positive IgG/IgM specimens detected by the Healgen® lateral flow rapid cassette nasopharyngeal swabs were also collected for confirmation COVID-19 by RT-PCR (Nucleic acid amplification technique or NAAT). Serum was stored at 2–8°C after centrifugation (Champion F-33D, AmpleScientific, 3300 RCF, 10 minutes) of the 3-4 mL blood containing 8.5 mL (16×100 mm) gel barrier SST vacutainer tubes. Nasopharyngeal swabs were placed in transport medium tubes and stored at 2 – 8°C prior to testing by RT-PCR. Processed lateral flow rapid cassettes were placed in biohazard bags and stored at 2–8°C.

### 2.5 Healgen® lateral flow rapid cassette kit procedure and result interpretation

Rapid cassettes and mini plastic dropper were removed from the sealed foil pouch and placed on a clean and level surface. Sample buffer was equilibrated at room temperature (15-30°C) prior to testing. Upon cleaning the finger stick collection site with the alcohol prep pad, whole blood was drawn by application of the 21G lancet, the 5 μL mini plastic dropper was used to apply whole blood specimen into the sample well (S) of the rapid cassette followed immediately by two drops (about 80 μL) of sample buffer to the buffer well (B) while avoiding air bubbles (**Fig 1a**). An additional drop of the sample buffer was added within 2 minutes if the red color did not move across the test window or if blood was still present in the specimen well (S). The result was read in ten minutes. Results interpretation after fifteen minutes was avoided.

**Fig 1.**
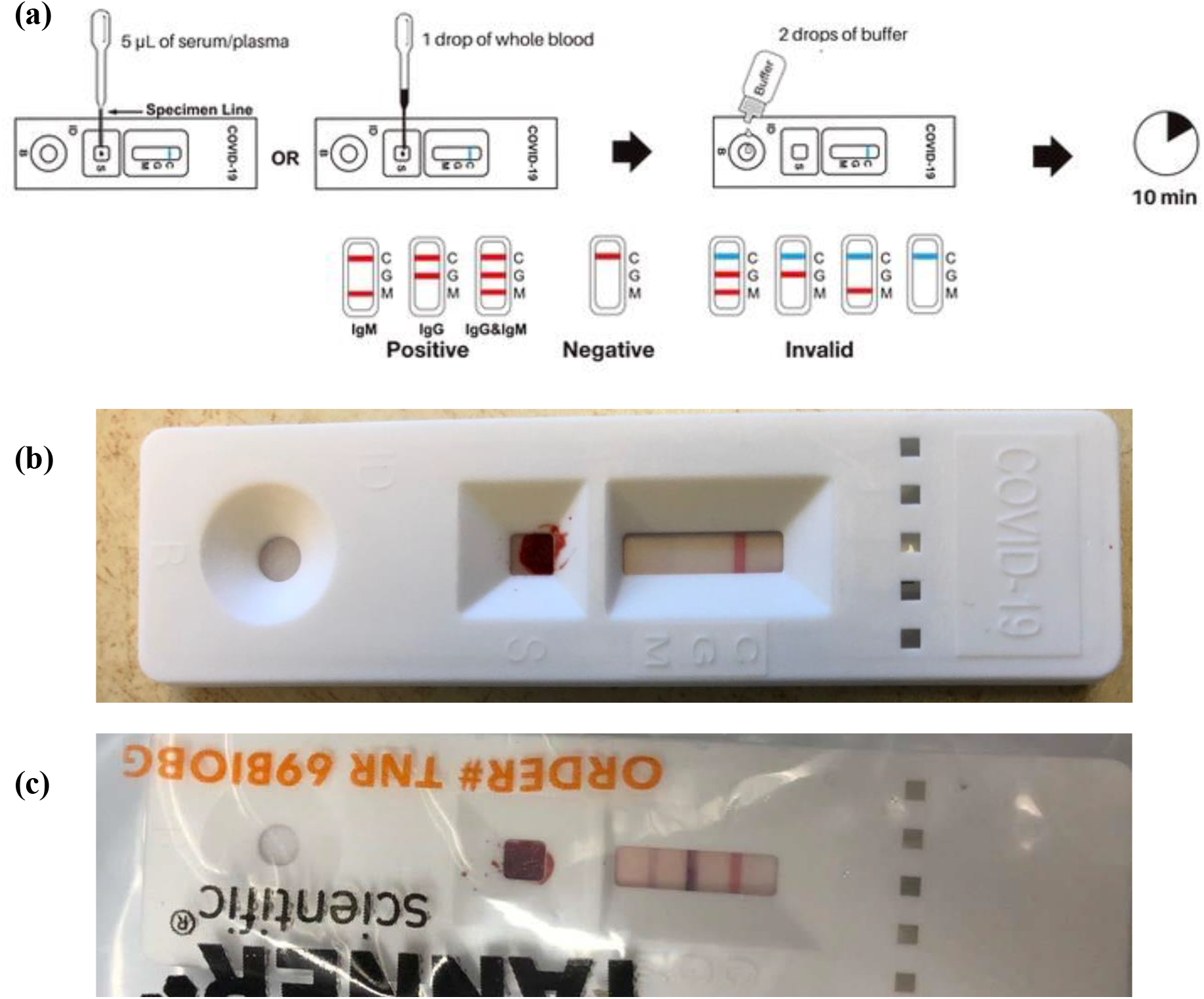
(a) Healgen® lateral flow rapid cassette kit procedure and result interpretation. (b) Negative result Healgen® rapid cassette. (c) IgG and IgM positive result Healgen® rapid cassette.

Results were interpreted as negative if the colored line in the control line region (C) changed from blue to red and no line appeared in the test line regions M or G (**Fig 1b**). IgM only positive was indicated if the colored line in the control line region (C) changed from blue to red, and a colored line appeared in test line region M. This test result indicates the presence of IgM anti-SARS-Cov-2 antibodies. If the colored line in the control line region (C) changed from blue to red, and a colored line appeared only in test line region G, the test was considered IgG positive - the test result indicates the presence of IgG anti-SARS-CoV-2 antibodies. For both IgG and IgM to be considered positive the colored line in the control line region (C) needed to have changed from blue to red, and two colored lines appeared in test line regions M and G. The test results indicate the presence of both IgM and IgG anti-SARS-CoV-2 antibodies (**Fig 1c**). An invalid result is indicated when the control line is partially red and failed to completely change from blue to red. Insufficient specimen volume or incorrect procedural techniques are the most likely reasons for control line failure.

### 2.6 EDI™ Novel Coronavirus COVID-19 IgG/IgM ELISA test procedure and result interpretation

Prior to use, all reagents were equilibrated at room temperature and reagents from different lots kept separate. ELISA wash concentrate was diluted to 1x working solution prior to use. An appropriate number of microwell strips were placed in a microwell plate holder to run 2 negative controls, 1 positive control and samples, once for IgG, once for IgM (for plate setup example see **Table 1**). For IgG, 10 μL of serum was diluted at a 1:100 dilution ratio with the COVID-19 IgG sample diluent and 100 μL of control solution (1 negative, 1 positive for IgG/IgM) was added into the designated microwells. For IgM 10 μL of serum sample was added into the designated microwells (no dilution required for IgM samples) followed by 100 μL of COVID-19 IgM sample diluent to the microplate wells. After gentle mixing (incubating microplate shaker, 250 rpm) the plate was sealed with aluminum adhesive foil. For IgG, incubation was performed at room temperature (20-25°C) for IgM at 37°C, each for 30 minutes. After incubation, the plate seal was removed and well contents aspirated. Each well was washed five times by dispensing 350 μL of diluted wash solution and aspiration using automated microplate washer (BIOTEK ELx-50).

**Table 1:** Plate setup for serum ELISA runs:

|  | STRIP1 | STRIP2 | STRIP3 |
| --- | --- | --- | --- |
| **A** | **Negative Control** | Sample 6 | Sample 14 |
| **B** | **Negative Control** | Sample 7 | Sample 15 |
| **C** | **Positive Control** | Sample 8 | Sample 16 |
| **D** | Sample 1 | Sample 9 | Sample 17 |
| **E** | Sample 2 | Sample 10 | Sample 18 |
| **F** | Sample 3 | Sample 11 | Sample 19 |
| **G** | Sample 4 | Sample 12 | Sample 20 |
| **H** | Sample 5 | Sample 13 | Sample 21 |

For IgG 100 μL HRP-labeled Anti-hIgG tracer antibody was added into the microwells, for IgM 100 μL of HRP-labeled COVID-19 antigen was added. The plates were covered with adhesive aluminum foil, mixed gently, and incubated at room temperature for IgG (20-25°C), for IgM at 37°C, each for 30 minutes. The plate seal was removed to aspirate the contents of each well. And each well washed 5 times by dispensing 350 μL of diluted wash solution into each well followed by aspiration of all well contents. Finally, 100 μL of ELISA HRP Substrate (Tetramethylbenzidine (TMB) with stabilized hydrogen peroxide) was added into the microwells, sealed and incubated during gently mixing at room temperature (20-25°C) for 20 minutes for both IgG and IgM plates. After seal removal 100 μL of stop solution was added into each of the microwells. Absorbance for each well was measured 20x at 450 nm within 10 minutes using the Safire2 microplate reader.

The average value of the absorbance of the negative control (xNC) was first determined to calculate the cutoffs using the following formulas for IgG and IgM (**Table 2**):

**Table 2:**
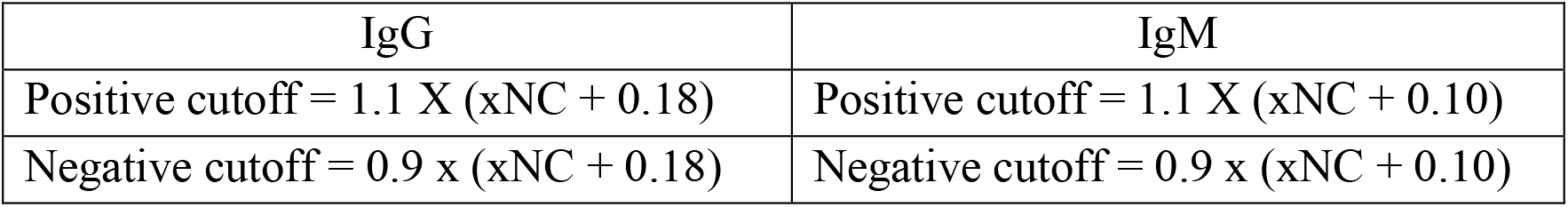
Formula to calculate cutoff values for EDI™ Novel Coronavirus COVID-19 IgG/IgM ELISA diagnostic kit:

| IgG | IgM |
| --- | --- |
| Positive cutoff = $1.1 \times (xNC + 0.18)$ | Positive cutoff = $1.1 \times (xNC + 0.10)$ |
| Negative cutoff = $0.9 \times (xNC + 0.18)$ | Negative cutoff = $0.9 \times (xNC + 0.10)$ |

Determination of result outcomes was based on cutoff values. If the measured values of the sample were equal or less than the negative cutoff, then the result was declared negative. If the measured values were equal or more than the positive cutoff value, then the result was declared positive. The results were considered borderline if the measured value was more than the negative cutoff but less then positive cutoff. Interpretation of the sample was performed by comparing the OD450 value to **Table 3**:

**Table 3:**
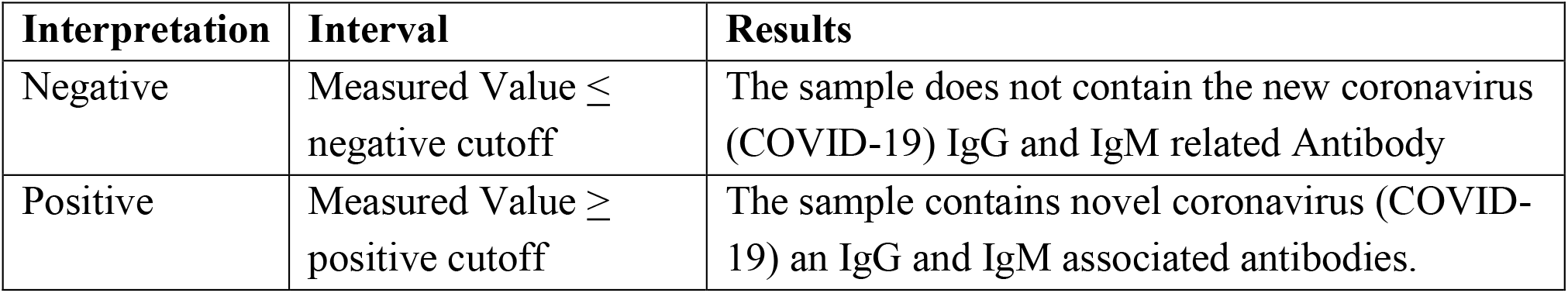
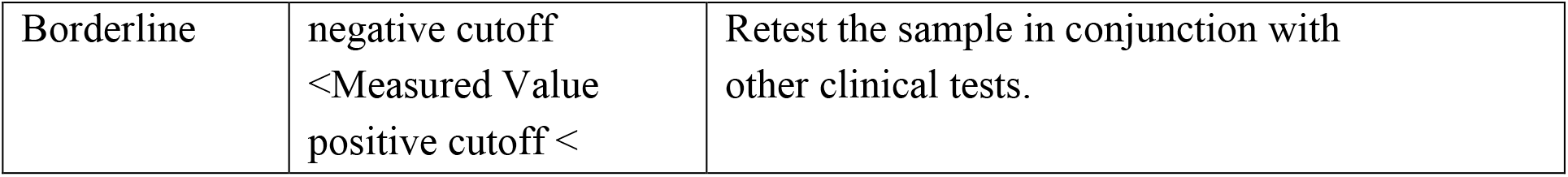
Interpretation of results based on sample OD_450_ values:

### 2.7 RT-PCR analysis by Luminex MAGPIX NxTAG® CoV Extended Panel

Of 3 mL nasopharyngeal transport medium, 300 µL was aliquoted to Hamilton Robotics tubes for RNA sample extraction using ZymoResearch Quick-DNA/RNA Viral MagBead™ Viral Kits (R2140/R2141). Bacteriophage MS2 internal control is included in the NxTAG® CoV Extended Panel Assay kit. This internal positive control was added to each sample prior to extraction. Briefly, extraction was performed by adding 10 μL of MS2 internal control reagent per 200 μL sample/quality control in a 96-deep well plate containing 400 μL Viral RNA Buffer. The plate was placed on a magnetic stand (strong-field magnetic stand or separator e.g., ZR-96 MagStand or Ambion 96 Magnetic Stand) until beads were pelleted. After 3 minutes, solution was aspirated and discarded by the Hamilton Robotics system. 500 μL MagBead DNA/RNA wash buffer 1 was added and mixed for 10 minutes on the Bioshake (1400 rpm). Beads were again pelleted on the magnetic stand and supernatant discarded. Additional wash cycles with 500 μL MagBead DNA/RNA wash buffer 2, 500 μL ethanol (95-100%) and 300 μL of ethanol (95-100%) were performed, with a transfer of the entire 300 μL sample suspension (beads and liquid) to an elution plate. Beads were pelleted on the magnetic stand supernatant discarded. Magnetic beads were dried for 10 minutes (beads will change in appearance from glossy black when still wet to a dull brown when fully dry). To elute DNA/RNA from the beads, the plate was removed from the magnetic stand 80 μL DNase/RNase-Free water added, the plate was mixed for 10 minutes on the Bioshake (1400 rpm) and transferred magnetic stand until beads were pelleted. 80 μL eluate was transferred to a new sample plate. 35 μL of the 80 μL eluate was added to the NxTAG® CoV Extended Panel Lyophilized Bead Reagents (LBRs) for an end volume of 50 μL and mixed by pipetting 10 times to resuspend the reaction reagents on an Eppendorf chilled PCR cooler block (stored in-20°C freezer prior to use). The plate was resealed using precut foil seals provided by Luminex® and transferred to the C1000 thermal cycler preheated to 42°C with a pre-heated lid at 110°C. The following PCR protocol was programmed into the thermal cycler (**Fig 2)**:

**Fig 2.**
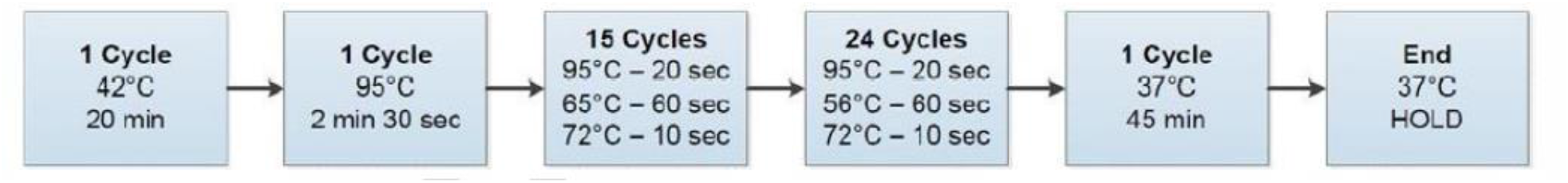
Thermal cycling parameters for the NxTAG® CoV Reaction Plate prior to analysis in the LUMINEX® MAGPIX® System.

The NxTAG® CoV Extended Panel (NxTAG CoV) incorporates multiplex Reverse Transcriptase Polymerase Chain Reaction (RT-PCR) with the Luminex® proprietary universal tag sorting system on the Luminex platform to detect SARS-CoV-2. Extracted total nucleic acid added to pre-plated, Lyophilized Bead Reagents (LBRs) was amplified via RT-PCR and the reaction product undergoes bead hybridization within the sealed reaction well. The hybridized, tagged beads were then sorted and read on the MAGPIX® instrument. The generated signals were analyzed using the NxTAG CoV Extended Panel Assay File for SYNCT™ Software, providing a reliable, qualitative call for SARS-CoV-2 viral target and internal controls within each reaction well.

### 2.8 Quality Controls

A procedural control is included in the Healgen® lateral flow rapid cassette test. A red line appearing in the control region (C) is the internal procedural control. It confirms sufficient specimen volume and correct procedural technique. Control standards are not supplied with the kit itself; however, it is recommended that positive and negative controls be tested as a good laboratory practice to confirm the test procedure and to verify proper test performance. Additional controls may be required according to guidelines or local, state, and/or federal regulations (such as 42 CFR 493.1256) or accrediting organizations. In this study pretested IgG or IgG, IgM positive serum control material and pretested IgG, IgM negative serum control material was pipetted onto 2 separate cassettes for every Healgen® lateral flow rapid cassette batch tested within 24 hours.

To assure the validity of the results of the EDI™ Novel Coronavirus COVID-19 IgG/IgM ELISA test kits, each assay must include both negative and positive controls. For IgG the average value of the absorbance of the negative control is less than 0.25, and the absorbance of the positive control is not less than 0.30. For IgM the average value of the absorbance of the negative control is less than 0.25, and the absorbance of the positive control is not less than 0.50.

Besides bacteriophage MS2 internal control included in the NxTAG® CoV Extended Panel Assay kit, SeraCare’s AccuPlex™ SARS-CoV-2 reference material kit for molecular (PCR) was used as positive reference material directed against the published CDC and WHO consensus sequences. SeraCare’s SARS-CoV-2 AccuPlex solution is non-infectious and replication deficient, enables safe handling of positive material, fully extractable with a real viral protein coat, and included negative reference material. Quality control material was stored at 2-8°C.

## 3. Results and Discussion

The Healgen® COVID-19 IgG/IgM Rapid Test Cassette was originally evaluated in China by testing a total of 191 plasma (K2EDTA) clinical samples and a study by the Frederick National Laboratory for Cancer Research (FNLCR) using frozen serum and plasma specimens, with 30 positives confirmed by RT-PCR and 80 negative specimens derived from collections before 2020 (n = 110). Alcala Labs has performed a similar study on clinical specimens, however this study of 75 specimen comparisons is unique in that it compares a whole blood finger-stick collection test method using the Healgen® rapid cassette to a simultaneous venipuncture draw serum collection measured by IgG/IgM ELISA, with RT-PCR confirmation of 8 out of 9 positives. Test results using the EUA-approved Healgen® COVID-19 IgG/IgM rapid cassette show that out of seventy-five total patient samples, sixty-four were negative for both IgG and IgM antibodies while nine were positive for both IgG and IgM antibodies. One sample turned out to be positive for IgG antibody only (**Table 4**).

**Table 4:** Summary results for the Healgen® COVID-19 IgG/IgM Rapid Test Cassette validated by testing whole blood finger-sticks versus simultaneous serum draws analyzed by an ELISA comparator method from individual donors (n = 75):

| Method | Comparator Method: (EDI™ Novel Coronavirus COVID-19 IgG/IgM ELISA) and RT-PCR for 8/9 positives |  |  |  |  |
| --- | --- | --- | --- | --- | --- |
|  | Antibody Positive |  |  | Negative | Total |
| Healgen® COVID-19 IgG/IgM Rapid Test Cassette | IgG+/IgM+ | IgG-/IgM+ | IgG+/IgM- | IgG-/IgM- |  |
| IgG+/IgM+ | 9 (**) |  | 1 |  | 10 |
| IgG-/IgM+ |  |  |  |  |  |
| IgG+/IgM- |  |  | 1 |  | 1 |
| Negative IgG-/IgM- |  |  | 1 (*) | 63 | 64 |
| **Subtotal** | 9 |  | 3 | 63 | 75 |

(\*) specimen was Negative (IgG-/IgM-) for whole blood, when serum was dripped onto the cassette directly from the venipuncture SST tube, the rapid cassette also showed no positive bands (IgG-/IgM-).
(\*\*) 8/9 of the IgG+/IgM+ cohort were confirmed to be COVID-19 positive by RT-PCR nasopharyngeal swab testing.

Out of the group of nine positive of the IgG+/IgM+ cohort eight were confirmed to be COVID-19 positive by RT-PCR (or Nucleic acid amplification technique = NAAT) swab testing. The same patients tested using the EDI™ Novel Coronavirus COVID-19 IgG/IgM ELISA test procedure with simultaneously provided venipuncture serum draws (SST) showed nine out of nine IgG+/IgM+ positives coincided with the Healgen® COVID-19 IgG/IgM rapid cassette results and confirmed one sample to be positive for IgG antibody only. Out of the 65 presumed negative for both IgG and IgM antibodies by the Healgen® COVID-19 IgG/IgM rapid cassette, the EDI™ Novel Coronavirus COVID-19 IgG/IgM ELISA test showed one specimen was positive for IgG only instead. The comparison to serum ELISA testing versus whole blood finger-stick testing using the Healgen® COVID-19 IgG/IgM rapid cassette resulted in the following statistics (**Table 5**):

**Table 5:** Summary statistics for the Healgen® COVID-19 IgG/IgM Rapid Test Cassette validated by testing whole blood finger-sticks versus simultaneous serum draws analyzed by an ELISA comparator method from individual donors (n = 75):

| Summary Statistics |  |  |
| --- | --- | --- |
| Measure | Estimate | 95% Confidence Interval |
| IgG Sensitivity/PPA | 100% (11/11) | (72.5%; 100%) |
| IgG Specificity/NPA | 98.5% (63/64) | (91.9%; 100%) |
| IgM Sensitivity/PPA | 90% (9/10) | (56.5%; 99.7%) |
| IgM Specificity/NPA | 100% (65/65) | (94.7%; 100%) |
| Combined Sensitivity/PPA | 100% (11/11) | (72.5%; 100%) |
| Combined Specificity/NPA | 98.5% (63/64) | (91.9%; 100%) |
| Combined PPV for prevalence = 5% | 77.8% |  |
| Combined NPV for prevalence = 5% | 100% |  |

Previously established sensitivity and specificity in the China study (**Table 6**) and the study by the Frederick National Laboratory for Cancer Research (FNLCR) [13] (**Table 7**) are summarized in **Tables 6** and **7**:

**Table 6:** Summary Results and Statistics for the Healgen® COVID-19 IgG/IgM Rapid Test Cassette validated by testing plasma (K2EDTA) clinical samples from individual patients exhibiting pneumonia, respiratory symptoms, and fever at two sites in China from January to mid - March 2020 (n = 191):

| Method | Comparator Method: RT-PCR |  |  |  |
| --- | --- | --- | --- | --- |
| Healgen® COVID-19 IgG/IgM Rapid Test Cassette |  | Positive | Negative | Total |
| Positive | IgG+/IgM+ | 78 | 0 | 78 |
|  | IgG-/IgM+ | 0 | 1 | 1 |
|  | IgG+/IgM- | 9 | 2 | 11 |
| Negative | IgG-/IgM- | 3 | 98 | 101 |
| **Subtotal** |  | 90 | 101 | 191 |
| **Summary Statistics** |  |  |  |  |
| **Measure** | **Estimate** |  | **95% Confidence Interval** |  |
| IgG Sensitivity/PPA | 96.7% (87/90) |  | (90.7%; 98.9%) |  |
| IgG Specificity/NPA | 98% (99/101) |  | (93.1%; 99.5%) |  |
| IgM Sensitivity/PPA | 86.7% (78/90) |  | (78.1%; 92.2%) |  |
| IgM Specificity/NPA | 99% (100/101) |  | (94.6%; 99.8%) |  |
| Combined Sensitivity/PPA | 96.7% (87/90) |  | (90.7%; 98.9%) |  |
| Combined Specificity/NPA | 97% (98/101) |  | (91.6%; 99%) |  |
| Combined PPV for prevalence = 5% | 62.9% |  |  |  |
| Combined NPV for prevalence = 5% | 99.8% |  |  |  |

**Table 7:** Summary Results and Statistics for the Healgen® COVID-19 IgG/IgM Rapid Test Cassette validated at the Frederick National Laboratory for Cancer Research (FNLCR) from frozen venipuncture serum and plasma samples (n = 110):

| **Method** | **Comparator Method: RT-PCR** |  |  | **Collected pre-2020** |  |  |
| --- | --- | --- | --- | --- | --- | --- |
|  | **Antibody Positive** |  |  | **Negative** | **Negative / HIV+** | **Total** |
| **Healgen® COVID-19 IgG/IgM Rapid Test Cassette** | **IgG+/IgM+** | **IgG-/IgM+** | **IgG+/IgM-** | **IgG-/IgM-** | **IgG-/IgM-** |  |
| IgG+/IgM+ | 29 |  |  |  |  | 29 |
| IgG-/IgM+ | 1 |  |  |  |  | 1 |
| IgG+/IgM- |  |  |  | 2 |  | 2 |
| IgG-/IgM- |  |  |  | 68 | 10 | 78 |
| **Subtotal** | 30 |  |  | 70 | 10 | 110 |
| **Summary Statistics** |  |  |  |  |  |  |
| **Measure** | **Estimate** |  | **95% Confidence Interval** |  |  |  |
| IgG Sensitivity/PPA | 96.7% (29/30) |  | (83.3%; 99.4%) |  |  |  |
| IgG Specificity/NPA | 97.5% (78/80) |  | (91.3%; 99.3%) |  |  |  |
| IgM Sensitivity/PPA | 100% (30/30) |  | (88.7%; 100%) |  |  |  |
| IgM Specificity/NPA | 100% (80/80) |  | (95.4%; 100%) |  |  |  |
| Combined Sensitivity/PPA | 100% (30/30) |  | (88.7%; 100%) |  |  |  |
| Combined Specificity/NPA | 97.5% (78/80) |  | (91.3%; 99.3%) |  |  |  |
| Combined PPV for prevalence = 5% | 67.8% |  |  |  |  |  |
| Combined NPV for prevalence = 5% | 100% |  |  |  |  |  |

The Healgen® COVID-19 IgG/IgM Rapid Test Cassette was originally evaluated using venipuncture serum or plasma specimens in China (n = 191 K2EDTA plasma specimens) or by the Frederick National Laboratory for Cancer Research (n = 110 serum or plasma specimens) with positives confirmed by RT-PCR. This study shows that a whole blood finger-stick collection versus venipuncture serum draws performs equally well considering combined sensitivity (100%) and 1% higher in combined specificity. Overall, the combined positive predictive value (PPV) at a presumed prevalence of 5% increased by 10% and the combined negative predictive value (PPV) at a presumed prevalence of 5% was maintained at 100% (**Table 8**):

**Table 8:** Summary Statistics comparison for the Healgen® COVID-19 IgG/IgM Rapid Test Cassette between the China study, FNLCR study and this study:

|  | **Summary Statistics** |  |  |  |
| --- | --- | --- | --- | --- |
|  | Combined Sensitivity/PPA | Combined Specificity/NPA | Combined PPV for prevalence = 5% | Combined NPV for prevalence = 5% |
| China study (n = 191) | 96.7% (87/90) | 97% (98/101) | 62.9% | 99.8% |
| FNLCR study (n = 110) | 100% (30/30) | 97.5% (78/80) | 67.8% | 100% |
| This study (n = 75) | 100% (11/11) | 98.5% (63/64) | 77.8% | 100% |

## 4. Conclusion

This study confirmed that utilizing the Healgen® COVID-19 IgG/IgM rapid cassette by applying whole blood finger-stick derived specimens instead of serum from a venipuncture draw maintains a diagnostic combined (overall) sensitivity of 100% and increases the diagnostic combined (overall) specificity to 98.5%. The negative predictive value (NPV) at 5% prevalence was maintained at 100% and the positive predictive value (PPV) increased by 10% to 77.8%. These values reflect an equal and higher performance (1% higher combined specificity, 10% PPV increase) than the performance characteristics described in the Healgen® COVID-19 IgG/IgM rapid test cassette (Whole Blood/Serum/Plasma) Instructions for Use (IFU) using whole blood/serum/plasma from venipuncture draws. This allows for application of the Healgen® COVID-19 IgG/IgM rapid test cassette in more remote or point-of-care settings where venipuncture draws may not be possible or affordable. A test performed with the Healgen® COVID-19 IgG/IgM rapid cassette from simple whole blood finger-stick collections is more efficient and equally reliable to serum ELISA testing or application of serum/plasma venipuncture draws.

## Data Availability

All data related to volunteer testing is summarized in the manuscript and was deidentified according to HIPAA guidelines.

## Author Contributions

Conceptualization, C.T. and D.J.S; methodology, I.B., E.B., C.T.; validation, I.B., E.B., C.T., M.J.C.M; formal analysis, R.S.Z., C.T.; Resources, D.J.S.; writing - original draft preparation, I.B., C.T., writing - review and editing, C.T.; supervision, D.J.S.; project administration, C.T.

## Funding

The authors received no specific funding for this work.

## Competing interests

The authors declare no competing interests.

## Acknowledgments

The authors would like to thank the San Diego Comprehensive Pain Management Clinic (SDCPMC) and associated staff for donor collections performed during this study.

## Notes

### Competing Interest Statement

The authors have declared no competing interest.

### Author Declarations

This study was approved by the Alcala Pharmaceutical Inc. Institutional Review Board (IORG0010127) in consideration of the Code of Ethics of the World Medical Association (Declaration of Helsinki). All collection activities occurred at the phlebotomy collection site at Alcala Testing and Analysis Services or the San Diego Comprehensive Pain Management Clinic (SDCPMC), 3703 Camino del Rio South, San Diego, CA, 92108 by certified phlebotomists or licensed vocational nurses. Serum ELISA testing and comparison to rapid cassette results were conducted from 06/01/2020 to 07/27/2020 at Alcala Testing and Analysis Services. All subjects were recruited via phone or e-mail announcements with phone follow-up or by word of mouth at the San Diego Comprehensive Pain Management Clinic, 3703 Camino del Rio South, San Diego, CA, 92108. Healthcare workers from the clinic also volunteered. Healthcare workers or volunteers in any setting could enroll without a prior COVID-19 test. All specimens derived from human subjects were de-identified of their health information as defined by Health Insurance Portability and Accountability Act (HIPAA) guidelines. Seventy-five volunteers took part in a whole blood finger-stick collection with immediate application to the Healgen COVID-19 IgG and IgM antibody rapid cassette and a simultaneous venipuncture draw of serum (SST tiger top tube) analyzed by the EDI Novel Coronavirus COVID-19 IgG/IgM ELISA test procedure.

